# Developing a deep learning model to predict the breast implant texture types with ultrasonography image: feasibility study

**DOI:** 10.1101/2024.03.18.24304460

**Authors:** Ho Heon Kim, Won Chan Jeong, Kyungran Pi, Angela Soeun Lee, Min Soo Kim, Hye Jin Kim, Jae Hong Kim

**Affiliations:** Department of Biomedical Informatics, Medical School of Yonsei University, Seoul; 3Billion, Inc, Seoul, South Korea; Qauntic EMBA; Korean Society of Breast Implant Research, Seoul, 03186, Republic of Korea; THE W Clinic, Seoul, Korea

**Author notes:** Address correspondence to: Jae Hong Kim, M.D., Current affiliation: The W Clinic, Address: 9F Kukdong B/D, 596 Gangnam-daero, Gangnam-gu, Seoul 06626 Korea.

## Abstract

**Introduction:** Breast implants, including textured variants, have been widely used in aesthetic and reconstructive mammoplasty. However, the textured type, which is one of the shell types of breast implants, has been identified as a possible carcinogenic factor for lymphoma, specifically breast implant-associated anaplastic large cell lymphoma (BIA-ALCL). Identifying the texture type of the implant is critical to the diagnosis of BIA-ALCL. However, distinguishing the shell type can be difficult due to human memory or loss of medical history. An alternative approach is to use ultrasonography, but this method also has limitations in quantitative assessment.

**Objective:** The objective of this study is to determine the feasibility of using a deep learning model to classify the textured shell type of breast implants and make robust predictions from ultrasonography images from heterogeneous sources.

**Methods:** A total of 19,502 breast implant images were retrospectively collected from heterogeneous sources, including images from both Canon (D1) and GE (D2), images of ruptured implants (D3), and images without implants (D4), as well as publicly available images (D5). The Canon (D1) images were trained using Resnet-50. The performance of the model on D1 was evaluated using stratified 5-fold cross-validation. Additionally, external validation was conducted using D2 and D5. The AUROC and PRAUC were calculated based on the contribution of the pixels with Grad-CAM. To identify the significant pixels for classification, we masked the pixels that contributed less than 10%, up to a maximum of 100%. To assess model robustness to uncertainty, Shannon entropy was calculated for four image groups: Canon (D1), GE (D2), ruptured implant (D3), and without implants (D5).

**Result:** The deep learning model achieved an average AUROC of 0.98 and a PRAUC of 0.88 in the Canon dataset (D1). For images captured with GE (D2), the model achieved an AUROC of 0.985 and a PRAUC of 0.748. Additionally, the model predicted an AUROC of 0.909 and a PRAUC of 0.958 for a dataset available online. For quantitative validation, this model maintained PRAUC up to 90% masking of less contributing pixels, and the remnant pixels located in breast shell layers. Furthermore, the prediction uncertainty increased in the following order: Canon (D1), GE (D2), ruptured implant (D3), no implant (D5) (0.066; 0072; 0.371; 0.777, respectively).

**Conclusion:** We have demonstrated the feasibility of using deep learning to predict the shell types of breast implants. With this approach, the textured shell types of breast implants can be quantified, supporting the first step in the diagnosis of BIA-ALCL.

## Introduction

Breast implants have been developed for aesthetic and reconstructive mammaplasty since 1962. The first textured breast implant was developed in 1968 to prevent capsular contracture after aesthetic or reconstructive implant-based mammaplasty [1,2]. Engraving and embossing types of textured implants have also been used in anatomical breast implants for natural shape. Since the first case of breast implant-associated anaplastic large cell lymphoma (BIA-ALCL) in 1997 by Dr. Keech, a total of 1264 cases of BIA-ALCL, including 59 deaths, have been reported by the Food and Drug Administration (FDA), according to a recent update as of June 30, 2023, [3,4]. Since the first case of BIA-ALCL, numerous investigations have been conducted to examine the etiology, prevalence rates, and clinical characteristics of BIA-ALCL.

Several studies have demonstrated that the topography of a textured breast implant shell surface is associated with BIA-ALCL [5–11]. Classified as a rare T-cell lymphoma, BIA-ALCL is nevertheless a significant concern in the context of breast augmentation and reconstruction surgeries, with documented cases of mortality. BIA-ALCL is often treatable when detected early, underscoring the critical importance of timely and accurate diagnosis [6,9–11]. In a real-world setting, identifying the inserted breast implant shell surface topography is the first step before diagnosing BIA-ALCL. If a patient has a history of primary aesthetic or reconstructive mammaplasty utilizing a smooth-type breast implant, there is generally no cause for concern regarding BIA-ALCL.

Nevertheless, a substantial number of patients may not be aware of the specific type of breast implant shell inserted during surgery over a long period of time, and medical records, especially within private clinics, may not be well preserved. Additionally, there are flaws in government policy in the regulation of medical devices in Korea [12]. As the medical community deepens its understanding of the complexities of this condition, the significance of diagnosing the surface topography of breast implant shells becomes increasingly apparent. Traditional diagnostic methods for assessing breast implants have relied heavily on subjective human evaluations. Mammograms and MRI, which are used to monitor breast implant-related complications, cannot identify the implant shell surface topography. Only ultrasonography can identify the inserted implant shell surface topography [13,14].

However, ultrasonography also has many limitations. The generalizability of the results may be limited due to the real-time and operator-dependent nature of ultrasonography, which may result in inter- and intra-observer variability. Although useful, to overcome these limitations of ultrasound, we conducted this study to determine whether it is useful in distinguishing breast implant shell surface topography using our algorithm. Additionally, the development of artificial intelligence (AI) programs aimed at accurately diagnosing breast implant shell surface topography holds promise in revolutionizing early detection and management strategies for BIA-ALCL. The development of AI algorithms aims to eliminate subjectivity and provide a more accurate and standardized approach to identifying the breast implant shell surface topography as textured and smooth type.

## Methods

### Study design

In this study, we retrospectively collected anonymous and de-identified medical records containing information on shell types of implants and ultrasonographic images, as well as demographic characteristics. We built multiple datasets as follows: 1) Canon dataset (D1), 2) GE dataset (D2), 3) ruptured implant dataset (D3), 4) no implant image dataset (D4), and publicly available dataset (D5). We used the Canon dataset (D1) for training, interval validation, and testing; GE and publicly available dataset for external validation. Additionally, the ruptured implant dataset and no implant images were used as out-of-distribution datasets to identify model interpretation.

First, the Canon and GE datasets (D1, D2) include the ultrasonography images with medical data generated from patients who underwent aesthetic or reconstructive implant-based mammaplasty without implant rupture at a single institution in South Korea. All patients underwent both breast cancer examination and ultrasonography-assisted examination at the institution. Ultrasonography assisted examination were conducted with an Aplio i600 (Canon Medical System, Otawara, Tochigi, Japan) system with a 7-18-MHz linear transducer, General Electric LOGIQ™ E10. We used these retrospective data confirmed by a surgeon between 31 August 2017 and 31 November 2022. We finally obtained the ultrasonography with medical data from 1,043 patients (Supplementary data 1). Multiple ultrasonography images from each patient were captured to assess the shell types of implants and saved in PACS rendered JPEG format. To rule out data leakage between train and test dataset, we retained unique images for model development by checking 128-bit MD5 hash algorithms. A breast surgeon with 14 years of breast implant ultrasonography experience labeled all the ultrasonographic images of shell surface topography.

In our problem, shell surface topography was divided into texture and smooth. The smooth type included micro-texture and nanotexture as conventional clinical classification [14]. Micro-texture and nanotexture types show almost the same shell surface topography with smooth type in high-resolution ultrasonography and light microscopy [14]. As some retrospective data were not stored as a DICOM (Digital Imaging and Communications in Medicine), we instead used only the centered PCAS-rendered image (Table 1) by discarding the top 12%, bottom 10%, and left and right 7% of pixels.

**Table 1.** Eligible ultrasonography dataset.

| Dataset name | Device | Objective | Shell integrity | Shell surface topography (N=19,502) |  |
| --- | --- | --- | --- | --- | --- |
|  |  |  |  | Texture | Smooth |
| Canon (D1) | Canon | Train, validation, test | Intact | 2,420 | 14,976 |
| GE (D2) | GE | External validation | Intact | 113 | 1,844 |
| Ruptured implant (D3) | Canon | OOD | Ruptured | 101 | 30 |
| Without implant (D4) | Canon | OOD | NA | NA (338) |  |
| Publicly available(D5) | Heterogenous | External validation | Intact | 11 | 7 |

Secondly, 131 ultrasonography images of the ruptured implant (D3) were collected using the Canon Aplio i600. Because of the damaged shell integration, the shell type of implant would be less easily identified from ultrasonography image. We used these images as out-of-distribution (OOD) dataset to identify the ability of model to estimate uncertainty of model for two types of shells. Thirdly, 338 ultrasonography images without implants were also captured, and used as OOD dataset to determine transparency of our model by estimating uncertainty (D4). Finally, we constructed a publicly available dataset for the external validation by searching for ultrasonography images using the following keywords: “breast implant ultrasound”, “breast implant ultrasonography” (D5).

### Model development

Convolutional neural networks were used by scaling the parameter sizes to achieve high performance. In this feasibility study, we used an off-the-shell convolution neural network (CNN) architecture originally designed for natural images, ResNet-50 composed of 50 layers as backbone [15]. To speed up the proof of concept, we chose a lightweight model instead of large CNN models such as Vision transformer or SwinTransformer, which require expensive computational costs with a large amount of data due to the lack of inductive bias [16,17]. We also trained our model by conducting transfer learning on pre-trained ResNet-50, which learned ImageNet classification. Then, we replaced the classifier layer of ResNet-50, which has 1,000-dimensional vector for multiclass into binary classifier layer for shell surface topology such as smooth, and texture. Weighted binary cross-entropy was used as the objective function for parameter optimization, which effectively trains the model by penalizing incorrect predictions of the minor class due to class imbalance between shell types (texture types being a minor class).

### Performance Evaluation

To transparently report the out model, we performed two types of validation: 1) stratified cross validation, 2) external validation with both GE dataset and publicly available ultrasonography image (D2, D3). First, stratified 5-fold cross-validation was conducted to identify generalized performance in a Canon dataset due to class imbalance between smooth and texture shells. Data split was performed by stratified random split ultrasonography images into training (60%), validation (20%), and test (20%) sets with shell surface topology labels. We evaluated the area under the curve (AUC) with different cut-off of receiver operating characteristic (ROC) curve and precision recall (PR) curve. Secondly, we conducted an external validation set to reduce latent bias with online acquired ultrasonography. We also identified AUC of both the ROC and PR curve with these data.

### Model interpretation: quantitative validation and uncertainty estimation

We used an explainable AI approach (XAI) to determine whether our model accurately classified the shell types of implants from the features of echogenicity, or layers from ultrasonography image and no other unexpected factors, by assessing the classification performance according to masking part of image. For the quantitative validation, we hypothesized that the important pixels of the image to distinguish the type of implant were on the layers of the implant. In addition, we also hypothesized that there would be no performance degradation if some non-layered pixels were erased. Therefore, we calculated the pixel importance using Grad-CAM, a method to quantify a pixel’s contribution to the classification [18]. Both AUROC and PRAUC were calculated by removing 10% of the least contributing pixels from the total number of pixels in the image. These pixels were replaced by zeros.

For the uncertainty estimation, we calculated the Shannon entropy to estimate the predictive uncertainty for each image in OOD [19,20]. Entropy ranges from 0 to 1, with a large value indicating greater predictive uncertainty. We also hypothesized that entropies in the ruptured implant dataset (D3) would be larger than those in the test set because our model trained on only ultrasonography image from a patient without damaged shell integrity. Furthermore, we hypothesized that the entropies in absent of breast implant images (D4) would be larger than those in the ruptured implant dataset (D3). Our model trained only ultrasonographic features from the implant image dataset to classify the shell types.

### Ethics

This retrospective study was approved by the Internal Institutional Review Board of the Korea National Institute of Bioethics Policy (IRB No. P01-202401-01-006), which waived the requirement for informed consent of medical records, including patients’ images and characteristics. All procedures described herein were performed by the 1964 Declaration of Helsinki and its later amendments or comparable ethical standards. None of the authors has a financial interest in any products, devices, or drugs mentioned in this manuscript.

## Results

### Classification performance for shell surface type

From stratified 5-fold cross validation, our model showed average AUROC of 0.98, and PRAUC of 0.88 in Canon dataset captured with Canon ultrasonography device (D1) (Figure 1-A, B). Overall, our model performed with an AUROC of 0.98 to 1.00, a PRAUC of 0.68 to 0.99 in Canon dataset (D1). Although the images were captured with GE ultrasonography device (D2), we identified our model showed AUROC of 0.985, and PRAUC of 0.748 (Figure 1-C, D). For publicly available dataset (D5), the model showed 0.909 of AUROC and 0.958 of PRAUC (Supplementary data 2).

**Figure 1.**
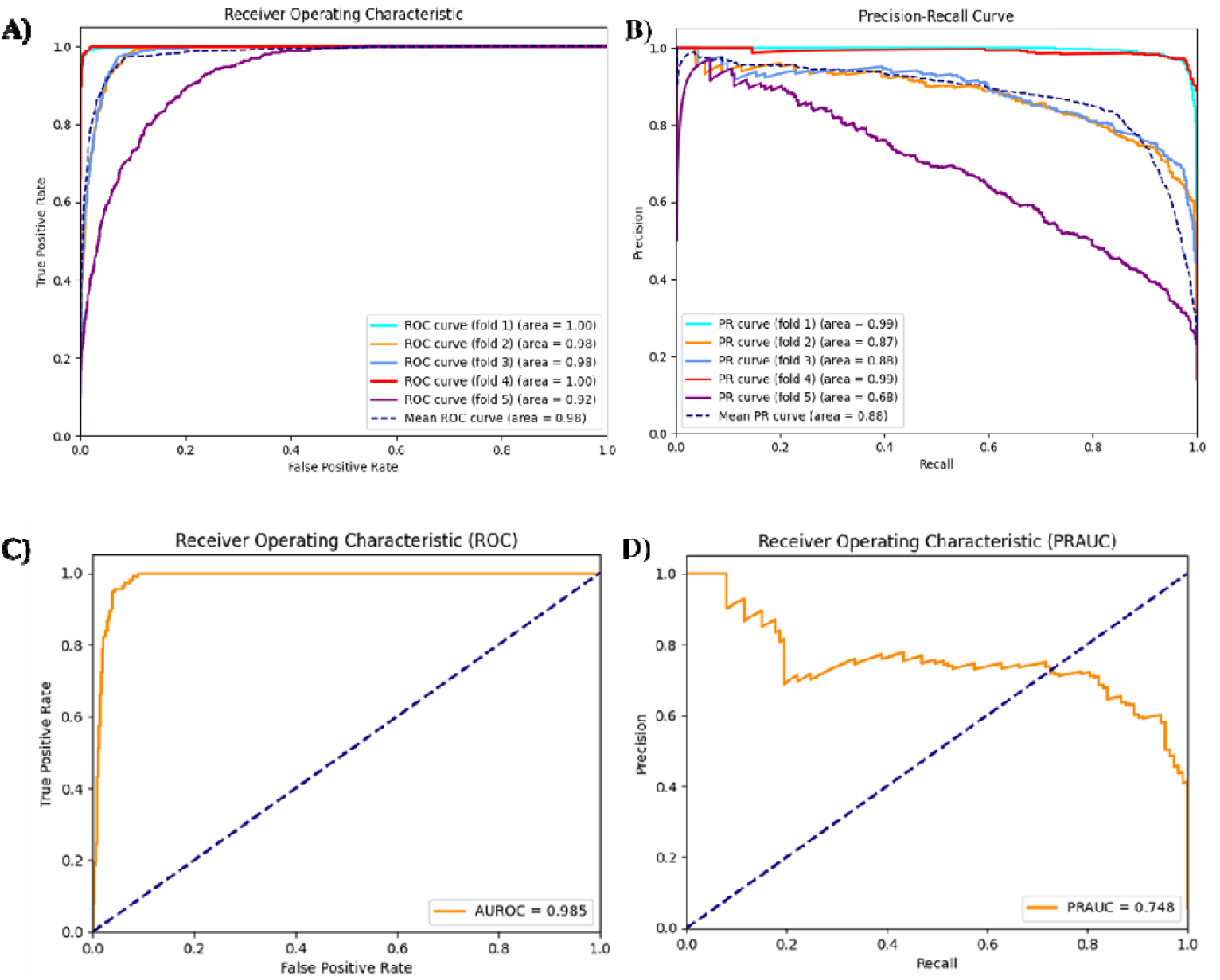
Shell types of classification performance. A) ROC curve for Canon dataset; B) PR curve for Canon dataset; C) ROC curve for GE dataset; D) PR curve for GE dataset;

### Quantitative validation

In the quantitative analysis to determine whether our model classifies ultrasonography images in accordance with medical knowledge, the model maintained an AUROC of 0.999 up to masking 90% of less contribution pixel to prediction, and showed AUROC of 0.997 when masking 100% of pixels. However, the PRAUC remained at 0.999 even after masking 90% of the pixels. After that, it decreased to 0.493 when all pixels were masked (Figure 2. Performance deterioration depends on masking non-contributing to prediction. A) Left. ROC curve for test dataset in Canon dataset; Right. PR curve for test dataset in Canon dataset; B) Texture shell implant prediction in Canon dataset (D1) by increasing the number of lower contributing pixels by 10%; C) Texture shell implant prediction in ruptured implant dataset (D3) by increasing the number of lower contributing pixels by 10%. For each individual case, the confidence for the texture shell type remained at 0.993 even when 80% of less contributing pixels were masked. At 90% of masking pixels, model confidence dropped into 0.968, and reached 0.497 when all of pixels were masked. Similarly, the model confidence for another case with texture shell types was maintained 0.994 until masking 80% of the pixels, decreased into 0.960 when masking 90% of pixels, dropped into 0.947 when masking 100% the of pixels (Figure 2-B, C).

**Figure 2.**
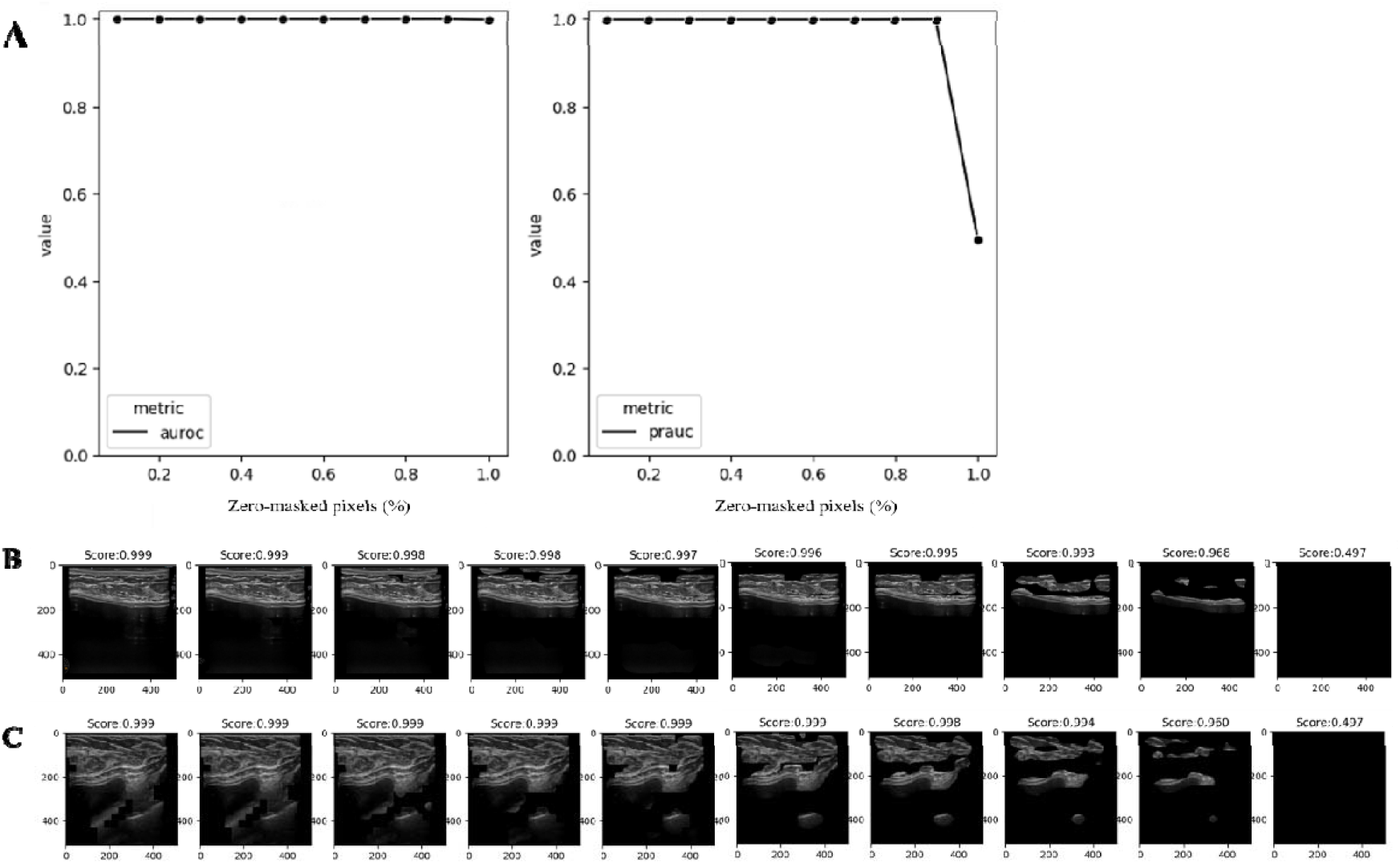
Performance deterioration depends on masking non-contributing to prediction. A) Left. ROC curve for test dataset in Canon dataset; Right. PR curve for test dataset in Canon dataset; B) Texture shell implant prediction in Canon dataset (D1) by increasing the number of lower contributing pixels by 10%; C) Texture shell implant prediction in ruptured implant dataset (D3) by increasing the number of lower contributing pixels by 10%.

### Uncertainty estimation

The model did not significantly produce lower entropies for the test dataset in Canon data (D1) than for the external validation set from GE ultrasonography device (mean [SD], 0.072[0.201] *vs* 0.066 [0.21]; *p*=0.350). However, the average entropy for ruptured implant images showed significantly high than for the test dataset in Canon data (mean [SD], 0.371 [0.318] *vs* 0.072[0.201]; *p*<.001). Moreover, model also predicted statistically significant high for the absent of implant image than for ruptured implant images (mean [SD], 0.777 [0.199] *vs* 0.371 [0.318]; *p*<.001) (Figure 3).

**Figure 3.**
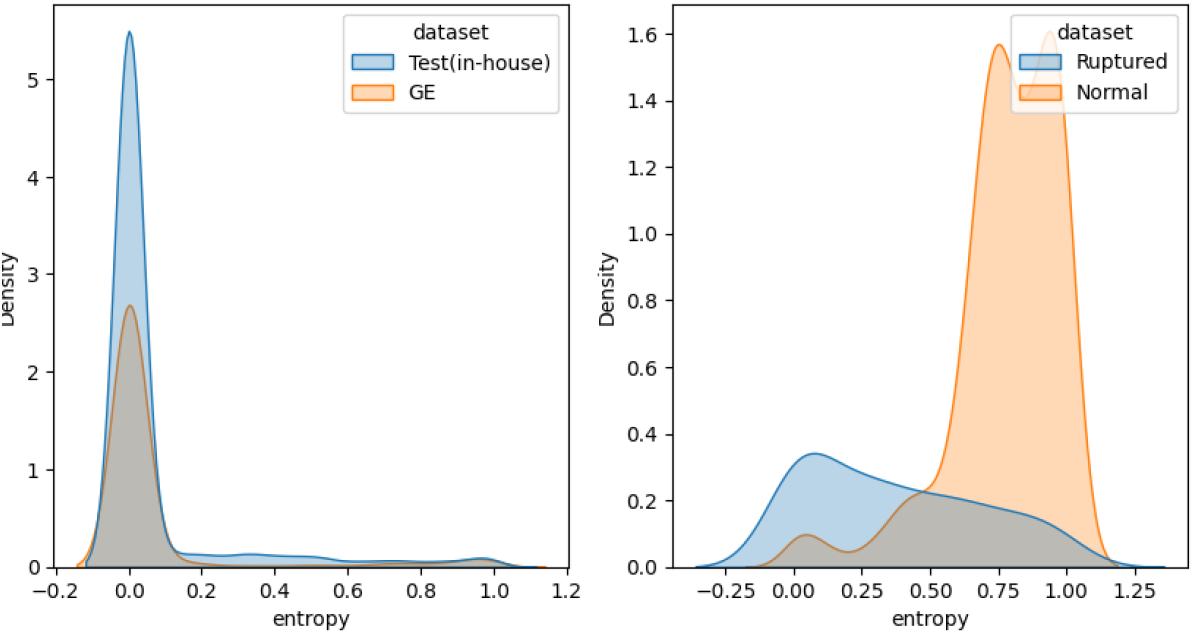
Entropy distribution of prediction from test in all datasets.

### Individual case review

For qualitative case review, we sampled two ultrasonography images; one from the test dataset (Canon, D1), and another from the ruptured implant datasets (D3), both captured by the same device. The model provided the model confidence of 0.998 as texture type for the texture shell type image. The Grad-CAM score for texture type showed the high value at shell in heatmap with Grad-CAM (white horizontal line in Figure 4-A). Also, for the image of ruptured texture shell implant, the model provided the model confidence of 0.664 as texture type. Although this score is higher than the classification threshold (0.5), it is lower than that of the intact texture shell type implant. However, the Grad-CAM showed high in the adjacent intact layer from ruptured shell area in the heatmap, despite the shell being ruptured due to shell tear (Figure 4-B).

**Figure 4.**
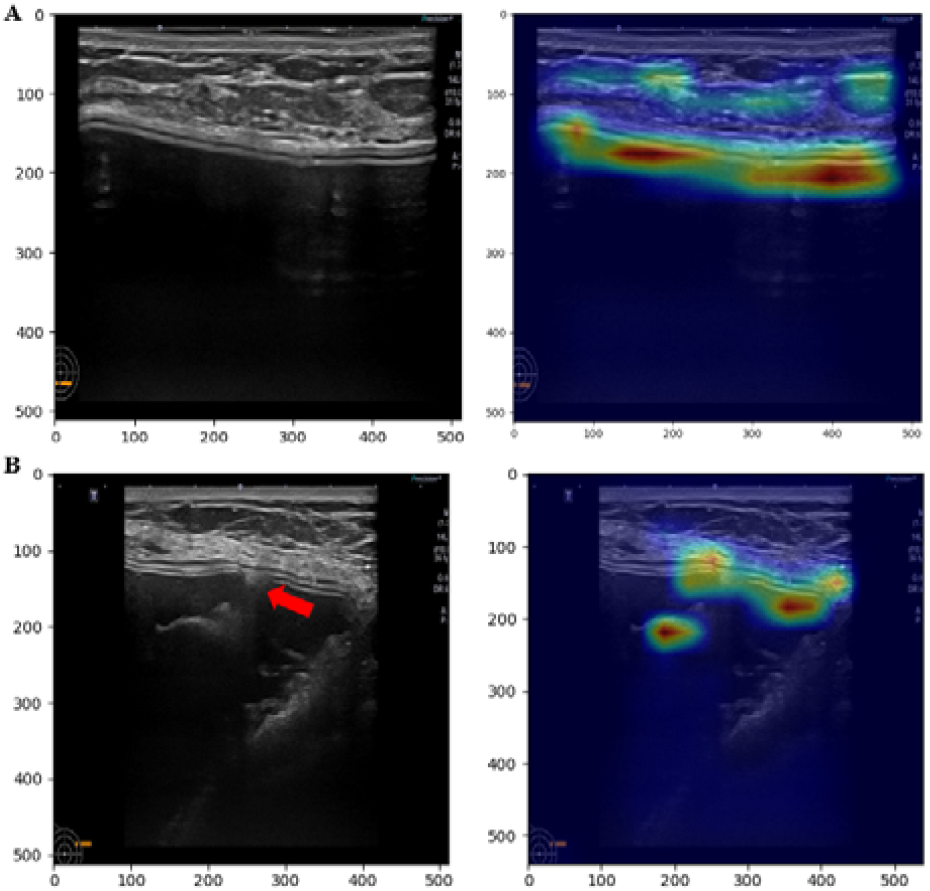
Preprocessed ultrasonography image with Grad-CAM for texture shell type prediction. Heatmap with Grad-CAM was bilinear interpolated to resize original image A) intact texture implant image captured with Canon ultrasonography device; B) damaged texture implant image captured with Canon ultrasonography device (the red arrow annotates shell tear).

## Discussions

### Principal Findings

The identification of breast implant shell types requires ultrasonographic examination, which can have inter- and intra-observer variability. Therefore, the generalizability of the results may be variable, leading to potentially missed diagnoses. However, there is no quantitative measurement, or classification method to distinguish the two shell types. This feasibility study demonstrates that deep learning can be used to quantitatively classify the shell type of breast implants. Also, this study supports the use of echogenicity from the shell layer of breast implants as an important region in classifying shell types. Furthermore, despite the use of different ultrasonography devices to capture images, our findings provide evidence that the deep learning model can classify the classes. Moreover, the model for ruptured breast implant ultrasonography images and ultrasonography images without absence of implant exhibit high uncertainty than the mages from intact shell type classification dataset, suggesting that the model could robustly quantify predictive uncertainty.

### Clinical application

Although there is a lack of standardized breast implant surface classification, ultrasonographic shell surface topographic images can divide the breast implant texture types into texture type and smoothness in high-resolution ultrasound [14]. Texture type shows roughness compared with smooth type in high-resolution ultrasonography (Supplementary data 3) [14]. Identifying the texture type of an inserted implant using ultrasonography instead of surgery is clinically important because the physician must consider the BIA-ALCL risk induced from the textured breast implant. As an extension of previous research on the feasibility of high-resolution ultrasonography for identifying the breast implant shell surface topography, this study was conducted to develop a deep-learning model to predict the breast implant texture types with ultrasonographic images [14]. In ultrasonography, smooth types include micro-texture and nanotexture types because micro-texture and nanotexture types show almost the same shell surface.

This method offers a promising way to classify breast implants with respect to the risk of BIA-ALCL, a condition that remains underexplored in current research. Given its rarity and the association of certain texture types with BIA-ALCL, accurate identification of texture type emerges as a critical determinant in risk assessment. The use of ultrasonography to identify texture type allows for straightforward identification on ultrasound images, often eliminating the need for additional testing. In addition, the use of deep learning models has the potential to assist patients undergoing breast augmentation or reconstruction, particularly in cases of implant rupture. Given the limited familiarity of radiologists and breast physicians with the diverse landscape of breast implants, including both manufacturers and shell types, the integration of AI in clinical contexts is proving invaluable.

### Reliable AI for clinical decision support

Reliable AI is essential for clinical decision support in the biomedical domain to avoid adverse patient outcomes[21,22]. This study includes the multiple experiments on different datasets such as difference device, and out-of-distribution dataset to explore the model transparency. In AI research for Radiology, it was found that the deep learning models often showed deteriorated performance in external validation [23]. The study reveal that deep learning models may be vulnerable to medical image from heterogenous sources due to unseen distribution. To eliminate biased evidence from these findings, we evaluated the model using ultrasonography image from heterogenous device (Figure 1). In addition, this study showed uncertainty in the model’s predictions, with the mean distribution being larger for images taken with the same device, images taken with different device, images with ruptured implants and images without implants (Figure 3). This can support that the deep learning model classify the shell type by learning ultrasonographic feature of breast implant shells. Further, for ruptured implant images, it is consistent with the medical field, where determining the shell type of a ruptured implant is difficult to observe due to the damaged surface of the implant (Figure 3). The entropies for ruptured implant images (D4) showed higher than intact implant images (D1, D2). This approach provides a model confidence that can help clinicians make decisions that reflect the uncertainty in the diagnosis when uncertainty is high, for example when the model consensus is close to 0.5, and more confident decisions when it is close to 1. Also, the important pixels can be provided to clinicians by conducting post-hoc analyses such as Grad-CAM or Score-CAM [18,24].

### Limitations

This study acknowledges several limitations that may introduce bias into interpretations. Primarily, the ultrasonography dataset did not represent all ultrasonography devices worldwide. Given the variability in the device resolution, configuration, and manufacturer, classification performance cannot be universally applied. To mitigate this, we performed internal and external validations on various ultrasound devices and incorporated out-of-distribution data to achieve less biased and more widely applicable results. In addition, the implant images collected did not include all types of shells used worldwide; rather we focused only on implants from eight manufacturers licensed by the Ministry of Food and Drug Safety of the Republic of Korea. As a result, a multicenter study spanning multiple nations and including images of common implants in each region would allow for more generalized interpretations of the results.

## Conclusion

The feasibility study presented demonstrates the potential of deep learning to accurately classify breast implant shell types from ultrasound images, addressing the current lack of standardized methods. Our findings underscore the importance of differentiating implant texture types, particularly for assessing the risk of BIA-ALCL. In addition, the adaptability of the deep learning model to account for imaging device variations and navigate prediction uncertainties opens promising avenues for robust AI-driven clinical decision support in the evaluation and management of breast implants.

## Data Availability

This dataset is available in https://github.com/4pygmalion/implant-shell-type.

## Notes

### Competing Interest Statement

The authors have declared no competing interest.

### Author Declarations

This retrospective study was approved by the Internal Institutional Review Board of the Korea National Institute of Bioethics Policy (IRB No. P01-202401-01-006), which waived the requirement for informed consent of medical records, including patients' images and characteristics

