## Supplementary data for "Developing a deep learning model to predict the breast implant texture types with ultrasonography image: feasibility study"

### Supplementary data 1

Supplementary table 1. Demographic characteristics of 1,043 patient.

| Variables | Values (mean±SD) |
| --- | --- |
| Age (years old) | 37.9±10.0 |
| Sex (male-to-female ratio) | 0:1043 |
| Height (cm) | 162.4±5.1 |
| Weight (kg) | 52.0±6.4 |
| BMI (kg/m^2^) | 19.7±2.2 |

Supplementary table 2. Characteristics of surgery and breast implant of 1,043 patients (2,068 breast implants).

| **Variables** | **Values (%)** |
| --- | --- |
| **Surgery** | |
| Bilateral  Unilateral | 1,050 (99.1)  18 (0.9) |
| **Purpose of surgery** | |
| Aesthetic  Reconstructive | 2,035 (98.4)  33 (1.6) |
| **Chamber** | |
| Single  Dual | 2,068 (100)  0 |
| **Fill material** | |
| Silicone  Saline | 2,004 (96.9)  64 (3.1) |
| **Shell type** |  |
| Texture  Smooth (including micro/nano-texture) | 403 (19.5)  1,665 (80.5) |
| **Manufacturer** | |
| HansBiomed Co., Ltd., Seoul, Korea    Groupe Sebbin SAS, Boissy-l’Aillerie, France    Mentor Worldwide LLC, Santa Barbara, CA, USA    Establishment Labs Holdings Inc., Alajuela, Costa Rica    Allergan plc, Dublin, Ireland    GC Aesthetics PLC, Apt Cedex, France    Polytech Health & Aesthetics, Dieburg, Germany    Silimed Inc., Rio de Janeiro, Brazil    Other | 594(28.7)  513(24.8)  257(12.4)  222(10.7)  215 (10.4)  85 (4.1)  56 (2.7)  22 (1.2)  104 (5.0) |
| **Surgery** | |
| Primary    Secondary  Rupture negative  Rupture present | 1,513 (73.2)    555 (26.8)  439 (21.2)  116 (5.6) |
| **Cause of Reoperation (n=265 case)** | **Values (%)** |
| Rupture  Baker III, IV capsular contracture  Dissatisfaction with shape  Fear of BIA-ALCL | 116 (43.8)  69 (26.0)  55 (20.8)  25 (9.4) |

### Supplementary data 2

For publicly available dataset (D5), we resized these ultrasonography images into 512 x 512 because this dataset has heterogenous image size. Then, we feed this dataset without any other preprocessing. The model showed 0.909 of AUROC and 0.958 of PRAUC. This dataset is available in <https://github.com/4pygmalion/implant-shell-type>.

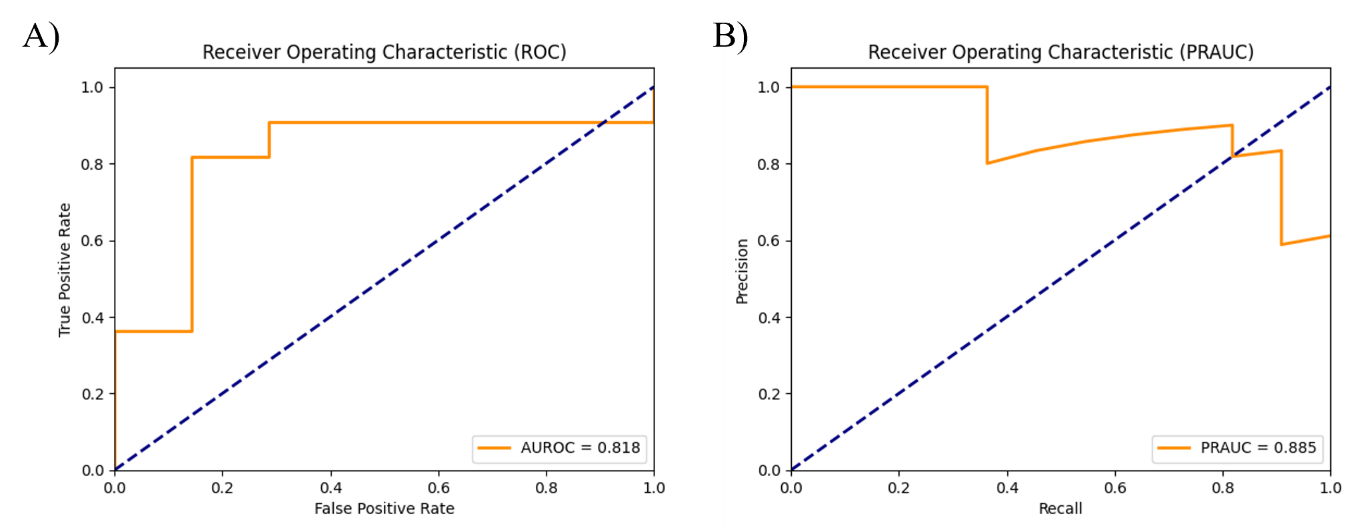

### Supplementary data 3

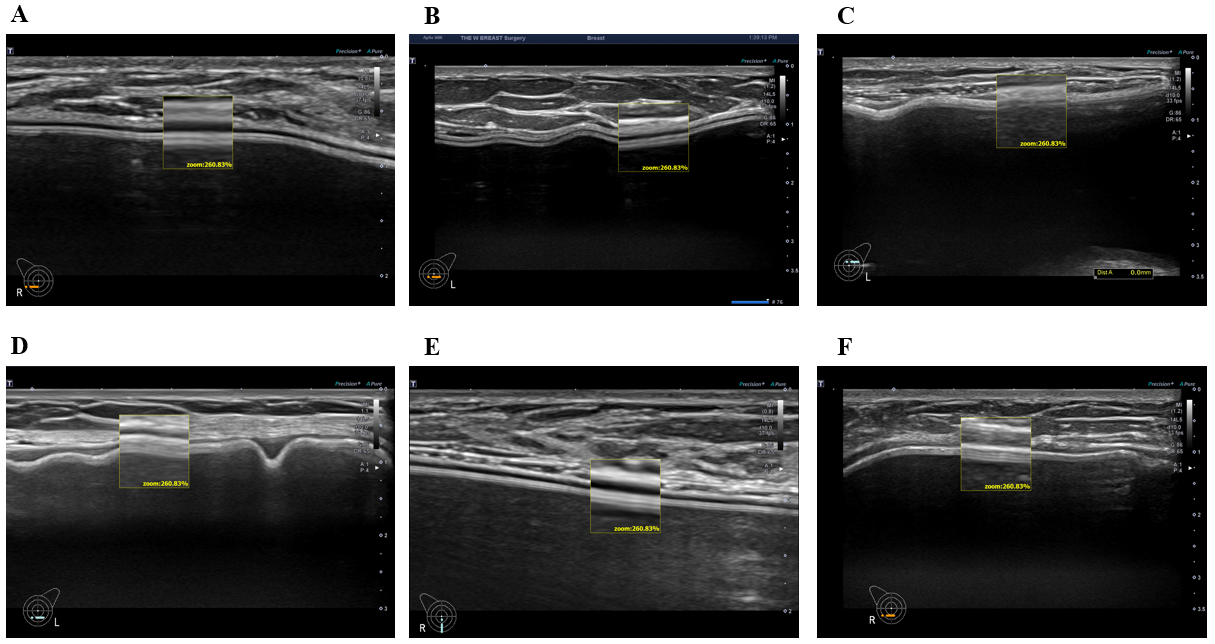

Supplementary figure 1. Example images of smooth type breast implants. A) Allergan smooth 3; B) Eurosilicone smooth; C) Mentor smooth 3; D) Sebbin smooth 2

Supplementary figure 2. Example images of texture type breast implants. A) Allergan Texture 5

B) Bellagle macro Texture 1; C) Eurosilicone Texture 1; D) Polytech Texture2; E) Sebbin Texture 1; F) Silimed texture 3

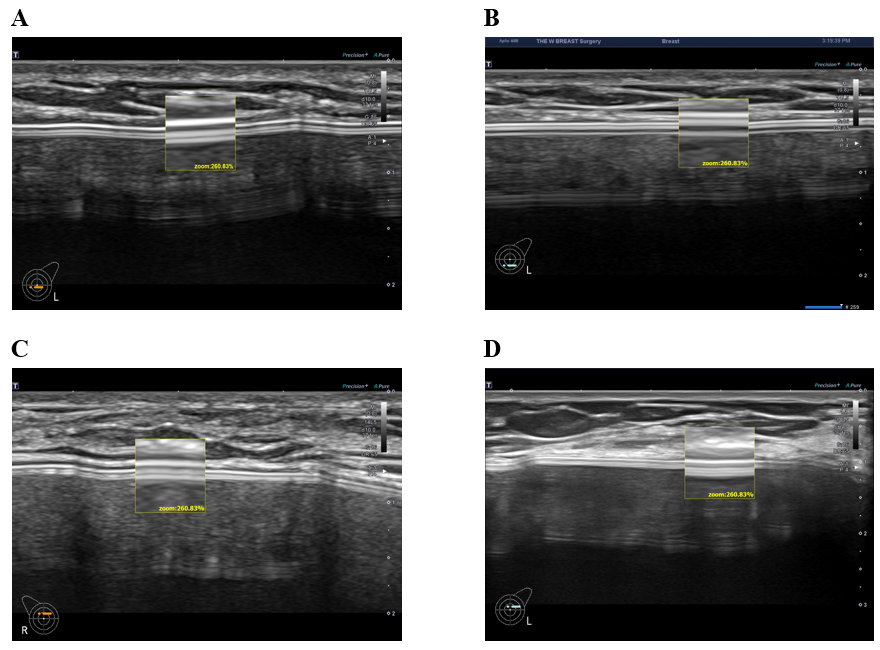

Supplementary figure 3. Example images of micro-texture type breast implants. A) Eurosilicone (GC Aesthetic); B) Hansbiomed Bellagel microtexture 3; C) Motiva microtexture; D) Sebbin microtexture

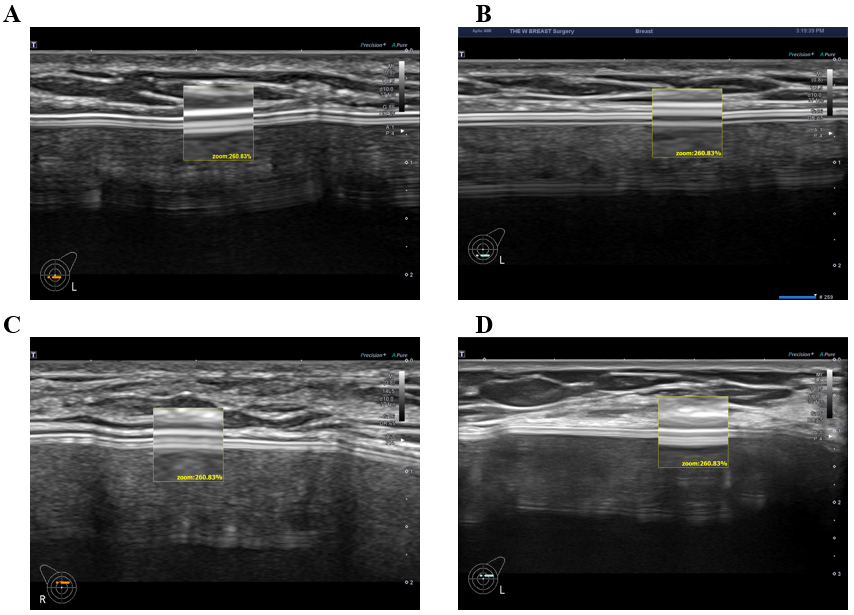
